# Demographic and Socio-Economic Factors, and Healthcare Resource Indicators Associated with the Rapid Spread of COVID-19 in Northern Italy: An Ecological Study

**DOI:** 10.1101/2020.04.25.20078311

**Authors:** Alessandra Buja, Matteo Paganini, Silvia Cocchio, Manuela Scioni, Vincenzo Rebba, Vincenzo Baldo

**Affiliations:** Department of Cardiologic, Vascular and Thoracic Sciences, and Public Health, University of Padova. Via Loredan, 18, 35131, Padova, Italy; Department of Biomedical Sciences, University of Padova. Via Marzolo, 3, 35131 Padova, Italy; Statistics Department, University of Padova. Via C. Battisti, 241, 35121, Padova, Italy; ‘Marco Fanno’ Department of Economics and Management, University of Padova. Via U. Bassi, 1, 35131, Padova, Italy

**Keywords:** COVID-19, pandemic, ecological studies, SARS-CoV-2

## Abstract

**Background:** COVID-19 rapidly escalated into a pandemic, threatening 213 countries, areas, and territories the world over. We aimed to identify potential province-level socioeconomic determinants of the virus’s dissemination, and explain between-province differences in the speed of its spread, based on data from 36 provinces of Northern Italy.

**Methods:** This is an ecological study. We included all confirmed cases of SARS-CoV-2 reported between February 24th and March 30th, 2020. For each province, we calculated the trend of contagion as the relative increase in the number of individuals infected between two time endpoints, assuming an exponential growth. Pearson’s test was used to correlate the trend of contagion with a set of healthcare-associated, economic, and demographic parameters by province. The virus’s spread was input as a dependent variable in a stepwise OLS regression model to test the association between rate of spread and province-level indicators.

**Findings:** Multivariate analysis showed that the spread of COVID-19 was correlated negatively with aging index (p-value=0.003), and positively with public transportation per capita (p-value=0.012), the % of private long-term care hospital beds and, to a lesser extent (p-value=0.070), the % of private acute care hospital beds (p-value=0.006).

**Interpretation:** Demographic and socioeconomic factors, and healthcare organization variables were found associated with a significant difference in the rate of COVID-19 spread in 36 provinces of Northern Italy. An aging population seemed to naturally contain social contacts. The availability of healthcare resources and their coordination could play an important part in spreading infection.

## BACKGROUND

The disease caused by the SARS-CoV-2 virus (COVID-19) first detected in Wuhan, China, in December 2019^1^ escalated rapidly into a pandemic,^2^ coming to threaten 213 countries, areas, and territories across the globe.^3^

Italy recorded its first imported cases in January 2020,^4^ and local cases emerged towards the end of February in Northern Italy, in the Lombardia and Veneto regions.^4,5^ Restrictions on international travel, domestic mobility, mass gatherings, and sporting events were gradually enacted by the Italian Government in an effort to contain the contagion,^4^ until the country’s complete lockdown on March 9^th^, 2020.^6^ Once implemented, these mitigation strategies (which sounded draconian to the population) proved effective in limiting the diffusion of COVID-19 to other parts of the country, but the infection continued to spread with different trends in Northern Italy.

Many demographic and socioeconomic factors, and healthcare organization variables can be associated with the variability of contagious disease propagation rates. It is important for public health systems to identify these factors and variables to produce solid evidence that can drive mitigation strategies. As well as having a direct influence on health outcomes in a given area, the findings of public health research also have important implications for ongoing global efforts to contain COVID-19. For example, a recent study found that even a small increase in long-term exposure to PM2.5 leads to a large increase in the virus-related death rate, with the magnitude of increase 20 times that observed for PM2.5 and all cause mortality.^7^

Besides environmental conditions, factors such as employment rates, population density, and healthcare resources may help to account for the observed differences in rapidity of COVID-19 propagation.

With this ecological study we aimed to identify potential province-level demographic, socioeconomic, and healthcare determinants of the COVID-19 pandemic’s dissemination, and to explain between-province differences in the rate of the disease’s spread, based on data from 36 provinces in four regions of Northern Italy.

## METHODS

### Context

The Italian National Health Service (NHS) was set up in 1978 with universal coverage, solidarity, human dignity, and human health as its guiding principles. It is regionally based, and organized at national, regional, and local levels. Under the Italian Constitution, central government controls the distribution of tax revenue for publicly-financed healthcare and decides on a national statutory benefits package of “essential levels of care” - offered to all residents in every region. The country’s 19 regions and two autonomous provinces are responsible for organizing and delivering healthcare services through local health units. Depending on the region, public funds are allocated by local health units to public hospitals and accredited private clinics. In 2017, there were approximately 213,700 hospital beds, including day hospital and day surgery beds: 170,000 in public hospitals (2.8 beds per 1,000 population, including 2.5/1,000 for acute care and 0.3/1,000 for long-stay patients), and 43,700 in accredited private clinics (0.7 beds per 1,000 population, including 0.4/1,000 for acute care and 0.3/1,000 for long-stay patients).^8^ Public hospitals are either managed directly by local health units and coordinated with other local health services such as preventive medicine departments and primary care districts, or they operate as semi-independent public enterprises. A prospective payment system based on diagnosis-related groups (DRG) operates across the country and accounts for most hospital revenue, but is generally not applied to hospitals run directly by local health units, where global budgets are common.

### Study design and data sources

In the present ecological study we included all confirmed cases of SARS-CoV-2 reported between February 24^th^ and March 30^th^, 2020, drawn from the Italian Civil Protection Department’s official data.^9^ This agency operates certified surveillance systems for the Italian government, with a two-stage testing procedure for SARS-CoV-2, one at local health unit level, and one at national laboratory level for confirmation purposes. All reported cases of COVID-19 infection met the laboratory criteria for confirmation established in the EU case definition procedure, which includes using RT-PCR.

We collected data on 36 provinces in four regions of Northern Italy – Piemonte, Lombardia, Veneto, and Emilia-Romagna – which have had the highest numbers of infections during the COVID-19 pandemic. For each province, we calculated the trend of contagion as the relative increase in the number of people infected between two time endpoints: T_0_, the day when the province first reached 10 cases, or as at February 2^nd^, 2020 (when the first data on the cases reported became available, whichever came first); and T_20_ (20 days after T_0_). Demographics were extracted from the most recently updated databases on the website of the Italian Institute of Statistics (ISTAT),^10^ and the ISTAT’s Health for All database.^11^ The number of sporting associations for each province was obtained from the registry of the Italian National Olympic Committee (CONI).^12^

### Data analysis

We modeled the disease’s spread assuming an exponential growth with the number of infections multiplying by a certain factor each day. Assuming no recoveries during the three-week period considered, the total number of people infected at the time t, y_t can be written as:

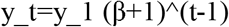

where y_1 is the number of cases on day 1 and β is the transmission factor.

Since we know the number of cases on day 1 and day 20, we can compute β_i, the transmission factor, for the i-th province as:

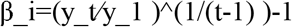

A descriptive analysis was conducted, calculating minimum, maximum, and mean values, to illustrate the spread of the disease. Pearson’s test was applied to correlate β_i with a set of healthcare-associated, economic, and demographic parameters at province level (see Table 1). β_i was input as a dependent variable in an OLS regression model to study the association between the rate of spread of COVID-19 and the variables that correlated with the dependent variable on a bivariate level with a p-value of less than 0.20. A stepwise selection procedure was used. A residuals analysis was performed to check the assumptions of the model. This analysis included testing for the residuals’ normal distribution (Shapiro-Wilk normality test) and independence (Durbin-Watson test).

**Table 1:**
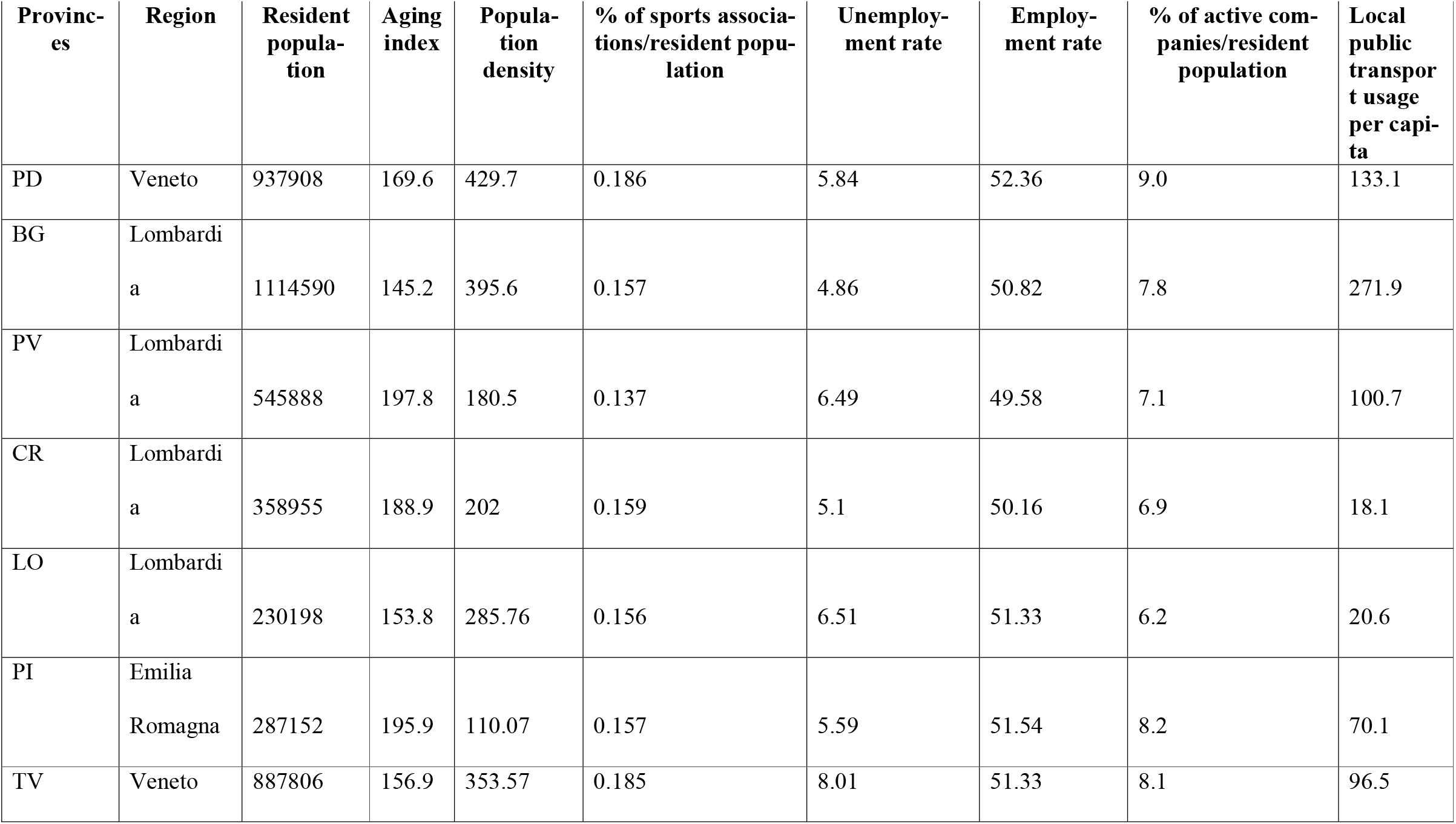
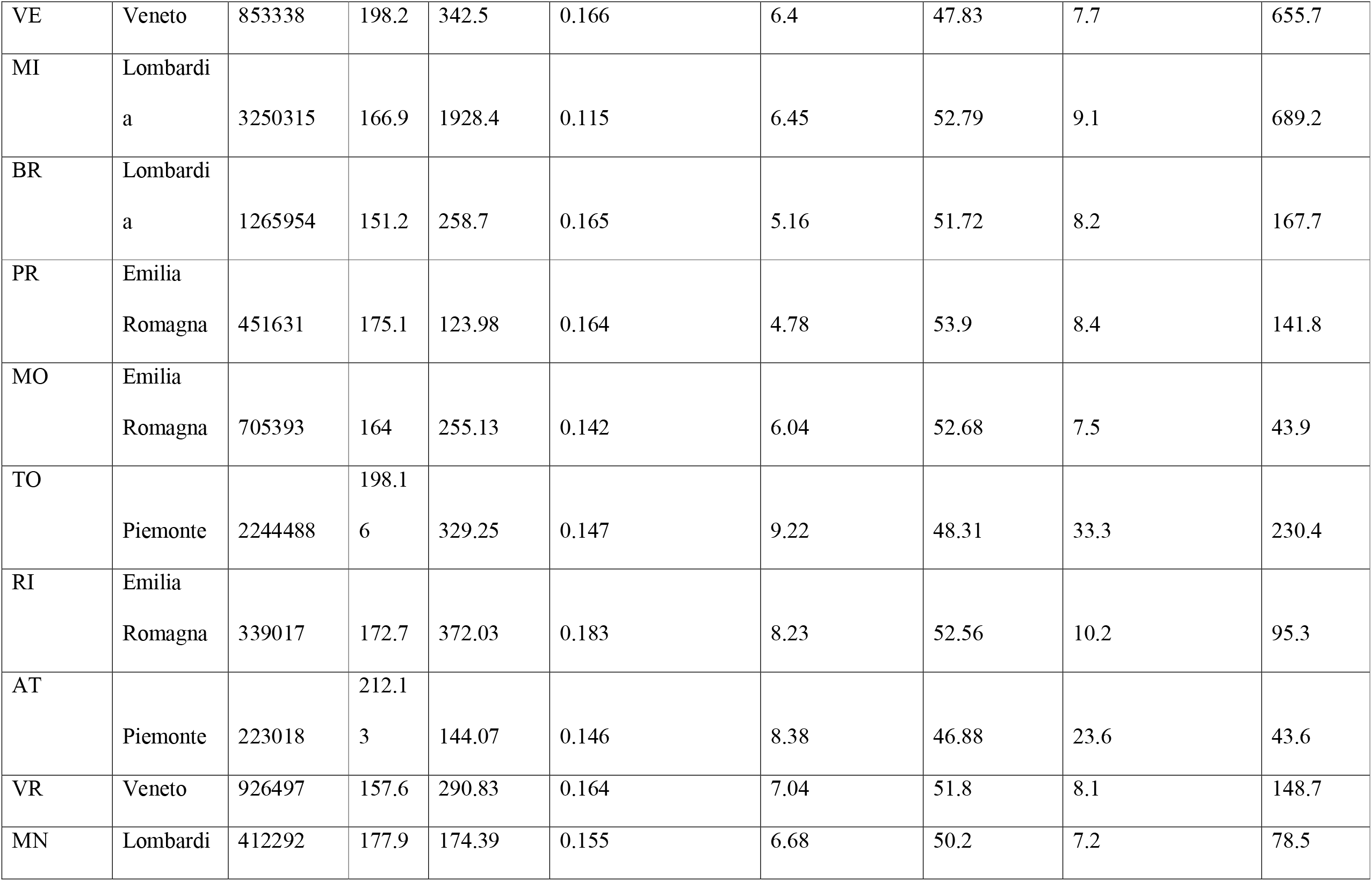
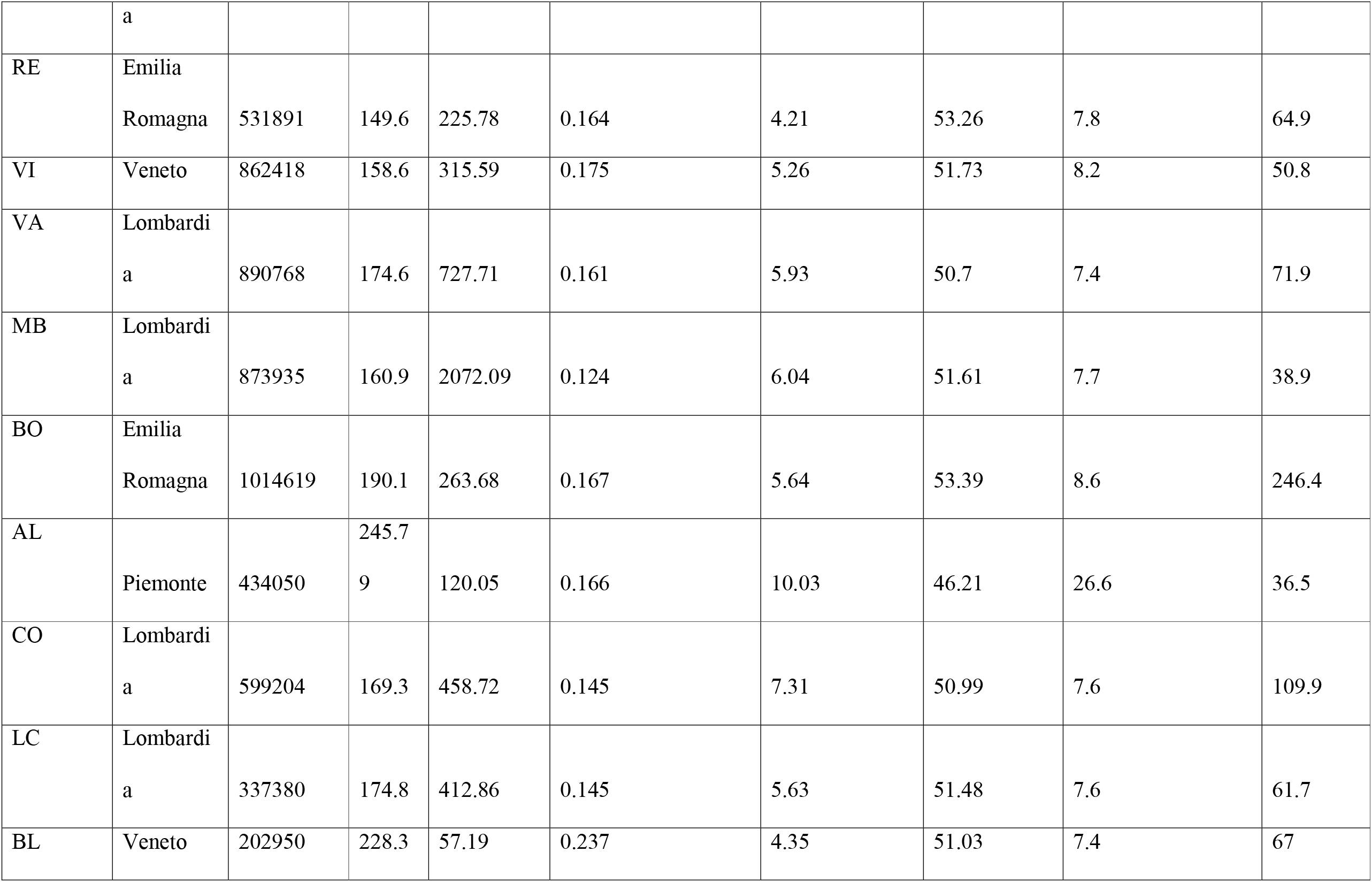
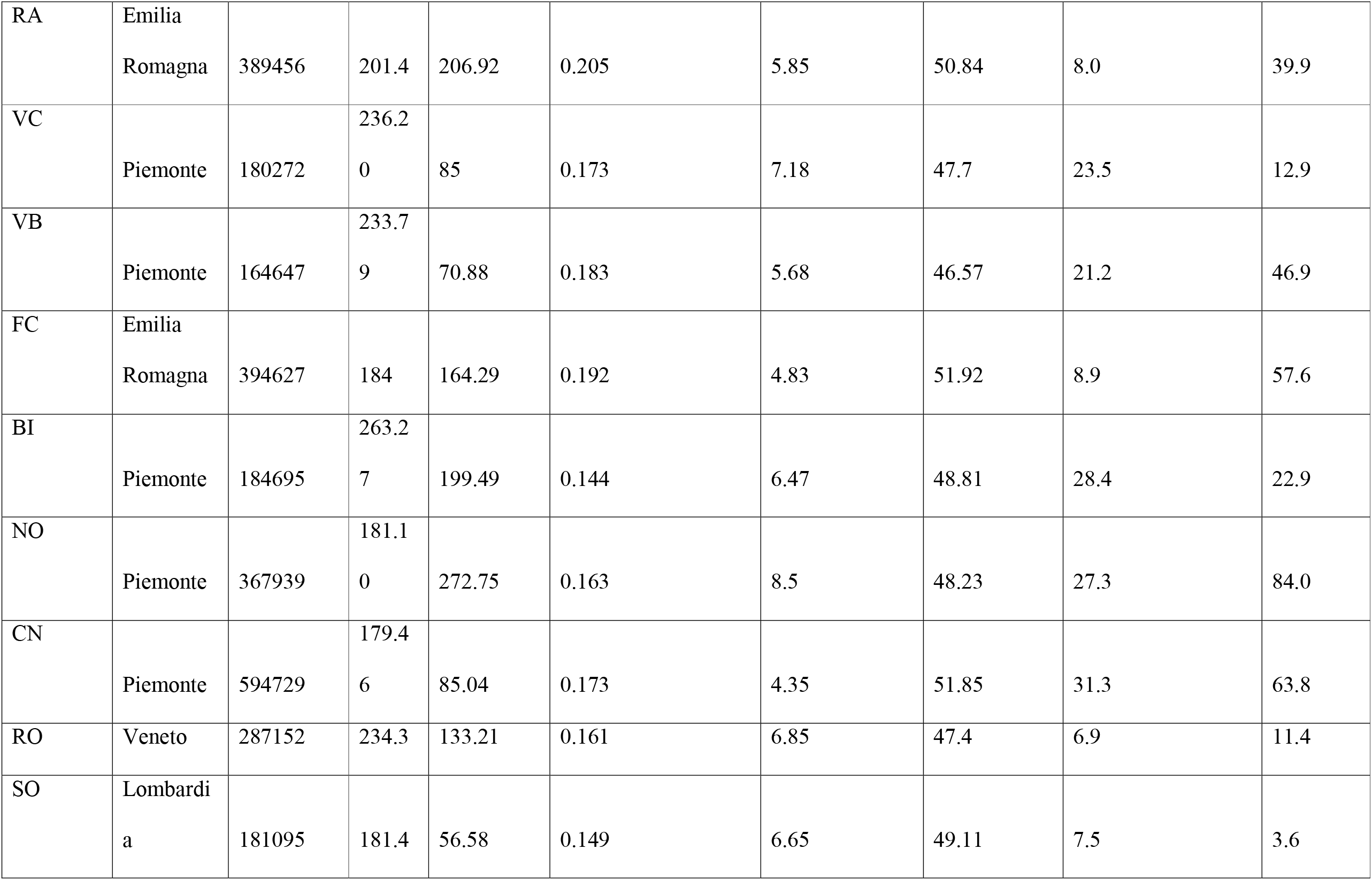
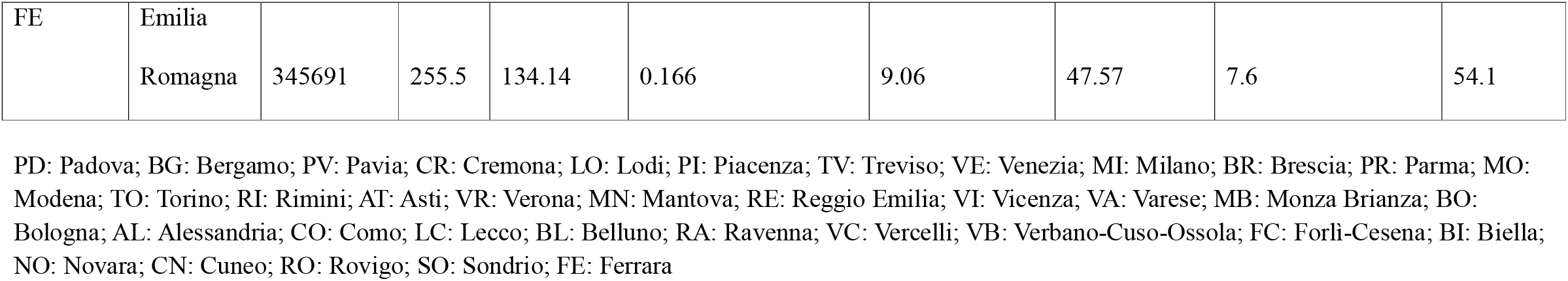
Environmental, health, economic, and population factors by province.

A sensitivity analysis was conducted, considering different dependent variables as rates (t1/t0) or incremental rates ([t1-t0]/t0), which produced the same results (data not shown). All data obtained were encoded in a master sheet using a Microsoft Office Excel spreadsheet (Version 2016, Microsoft Corporation, Redmond, WA). Data analysis was done with Stata 14 statistical software (Stata Corp LP, College Station, TX) and R Studio Integrated Development for R (RStudio, Inc., Boston, MA).

## RESULTS

Table 2 shows that the increase in the numbers of cases in 20 days varied considerably among the various provinces from a minimum of 115 cases in the province of Rovigo to a maximum of 3398 cases in Bergamo, with an average growth of 852 cases per province in 20 days. The rate of this increase was lowest in the province of Asti (4.5%) and highest in Brescia (330%), with an average over all provinces of 59.9%.

**Table 2:** Spread of COVID-19 (difference in cases, rate increase, incremental rate increase, and transmission rate β by province.

|  | **Table 2: Spread of COVID-19 (difference in cases, rate increase, incremental rate increase, and transmission rate $\beta$ by province** |  |  |  |  |  |  |  |
| --- | --- | --- | --- | --- | --- | --- | --- | --- |
| **Provinces** | **T0** | **Number of cases at T0** | **T1** | **Number of cases at T1** | **Difference in cases** | **Rate of increase in cases (T1/T0)** | **Incremental rate (T1-T0)/T1** | **$\beta$ (see Methods for calculation)** |
| **PD** | 25/02 | 30 | 15/03 | 658 | 628 | 21.9 | 20.9 | 0.176 |
| **BG** | 25/02 | 18 | 15/03 | 3416 | 3398 | 189.8 | 188.8 | 0.318 |
| **PV** | 25/02 | 27 | 15/03 | 722 | 695 | 26.7 | 25.7 | 0.189 |
| **CR** | 25/02 | 53 | 15/03 | 1792 | 1739 | 33.8 | 32.8 | 0.204 |
| **LO** | 25/02 | 125 | 15/03 | 1320 | 1195 | 10.6 | 9.6 | 0.132 |
| **PI** | 25/02 | 18 | 15/03 | 1012 | 994 | 56.2 | 55.2 | 0.236 |
| **TV** | 27/02 | 22 | 17/03 | 502 | 480 | 22.8 | 21.8 | 0.179 |
| **VE** | 27/02 | 14 | 17/03 | 378 | 364 | 27.0 | 26.0 | 0.189 |
| **MI** | 27/02 | 15 | 17/03 | 2326 | 2311 | 155.1 | 154.1 | 0.304 |
| **BR** | 27/02 | 10 | 17/03 | 3300 | 3290 | 330.0 | 329.0 | 0.357 |
| **PR** | 27/02 | 10 | 17/03 | 800 | 790 | 80.0 | 79.0 | 0.259 |
| **MO** | 27/02 | 18 | 17/03 | 460 | 442 | 25.6 | 24.6 | 0.186 |
| **TO** | 28/02 | 11 | 17/03 | 749 | 738 | 68.1 | 67.1 | 0.249 |
| **RI** | 29/02 | 15 | 19/03 | 691 | 676 | 46.1 | 45.1 | 0.223 |
| **AT** | 02/03 | 37 | 21/03 | 166 | 129 | 4.5 | 3.5 | 0.082 |
| **VR** | 03/03 | 17 | 22/03 | 1046 | 1029 | 61.5 | 60.5 | 0.242 |
| **MN** | 03/03 | 15 | 22/03 | 905 | 890 | 60.3 | 59.3 | 0.241 |
| **RE** | 03/03 | 14 | 22/03 | 1167 | 1153 | 83.4 | 82.4 | 0.262 |
| **VI** | 04/03 | 10 | 23/03 | 691 | 681 | 69.1 | 68.1 | 0.25 |
| **VA** | 04/03 | 11 | 23/03 | 421 | 410 | 38.3 | 37.3 | 0.211 |
| **MB** | 04/03 | 11 | 23/03 | 1130 | 1119 | 102.7 | 101.7 | 0.276 |
| **BO** | 04/03 | 11 | 23/03 | 833 | 822 | 75.7 | 74.7 | 0.256 |
| **AL** | 04/03 | 16 | 23/03 | 817 | 801 | 51.1 | 50.1 | 0.23 |
| **CO** | 05/03 | 11 | 24/03 | 635 | 624 | 57.7 | 56.7 | 0.238 |
| **LC** | 06/03 | 11 | 25/03 | 1076 | 1065 | 97.8 | 96.8 | 0.273 |
| **BL** | 07/03 | 11 | 26/03 | 313 | 302 | 28.5 | 27.5 | 0.193 |
| **RA** | 07/03 | 10 | 26/03 | 451 | 441 | 45.1 | 44.1 | 0.222 |
| **VC** | 07/03 | 10 | 26/03 | 336 | 326 | 33.6 | 32.6 | 0.203 |
| **VB** | 07/03 | 10 | 26/03 | 255 | 245 | 25.5 | 24.5 | 0.186 |
| **FC** | 08/03 | 15 | 27/03 | 580 | 565 | 38.7 | 37.7 | 0.212 |
| **BI** | 08/03 | 19 | 27/03 | 367 | 348 | 19.3 | 18.3 | 0.169 |
| **NO** | 08/03 | 13 | 27/03 | 609 | 596 | 46.8 | 45.8 | 0.224 |
| **CN** | 09/03 | 11 | 28/03 | 558 | 547 | 50.7 | 49.7 | 0.23 |
| **RO** | 10/03 | 10 | 29/03 | 125 | 115 | 12.5 | 11.5 | 0.142 |
| **SO** | 11/03 | 13 | 30/03 | 446 | 433 | 34.3 | 33.3 | 0.205 |
| **FE** | 11/03 | 12 | 30/03 | 306 | 294 | 25.5 | 24.5 | 0.186 |
| **MIN** |  | **10** |  | **125** | **115** | **4.5** | **11.5** | **0.082** |
| **Max** |  | **125** |  | **3416** | **3398** | **330** | **27.184** | **0.357** |
| **Mean** |  | **19** |  | **871.1** | **852.1** | **59.9** | **58.9** | **0.221** |

The increase in the numbers of cases in 20 days varied considerably among the various provinces.
T0 is the date of first details or 25.2. or report of 10th case. T1: 20 days after T0. PD: Padova; BG: Bergamo; PV: Pavia; CR: Cremona; LO: Lodi; PI: Piacenza; TV: Treviso; VE: Venezia; MI: Milano; BR: Brescia; PR: Parma; MO: Modena; TO: Torino; RI: Rimini; AT: Asti; VR: Verona; MN: Mantova; RE: Reggio Emilia; VI: Vicenza; VA: Varese; MB: Monza Brianza; BO: Bologna; AL: Alessandria; CO: Como; LC: Lecco; BL: Belluno; RA: Ravenna; VC: Vercelli; VB: Verbano-Cuso-Ossola; FC: Forlì-Cesena; BI: Biella; NO: Novara; CN: Cuneo; RO: Rovigo; SO: Sondrio; FE: Ferrara.

The scatter plot (Figure 1) shows significant correlations between the increase in transmission and several demographic, socioeconomic, and healthcare variables. A significant negative correlation emerged between increment β and population aging rates, while a significant positive correlation was found with employment rates, public transportation rates, in-house density, population density, and the proportions of private acute and long-term care beds in clinics and nursing homes.

**Figure 1:**
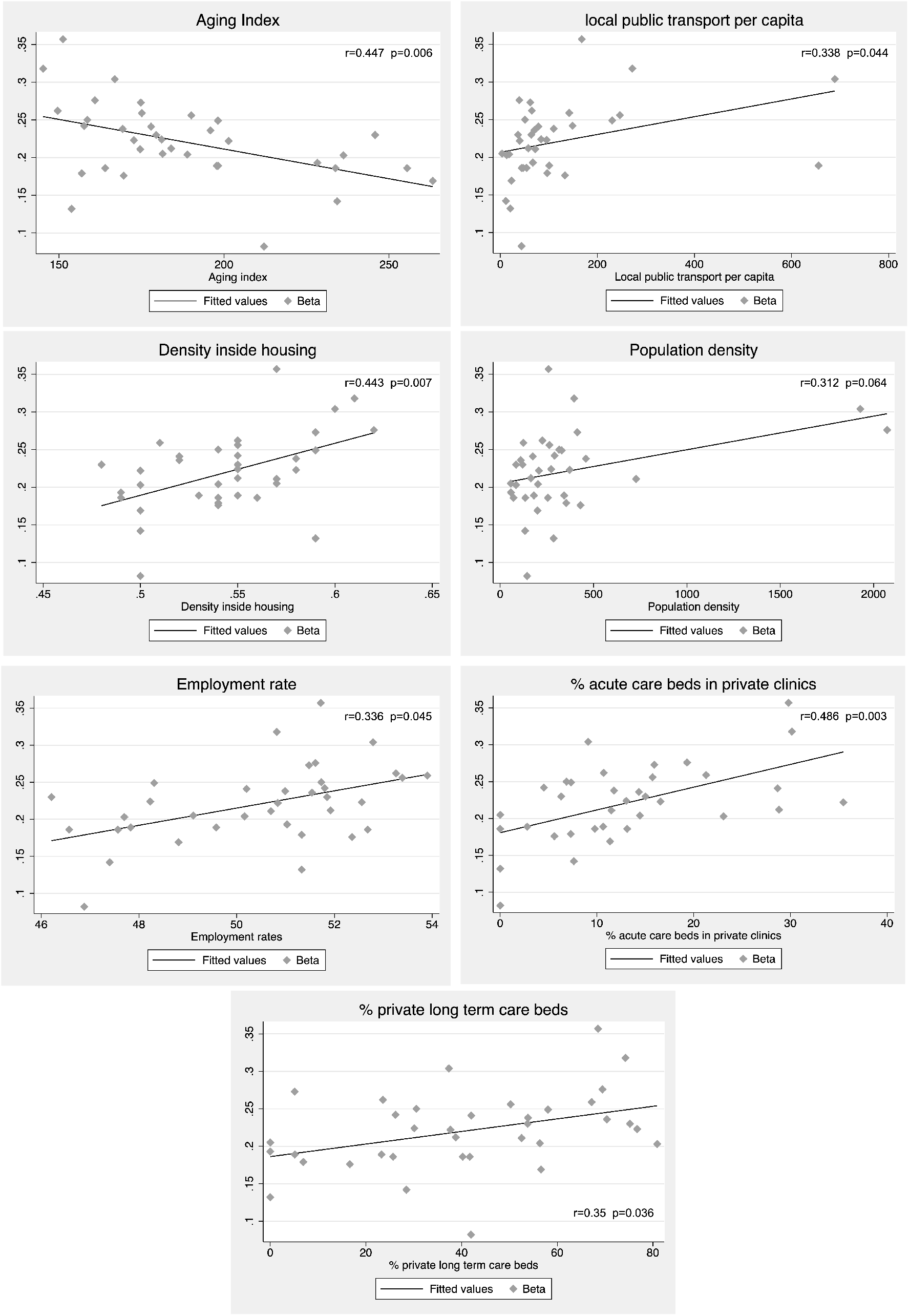
Significant correlations between β increment and · socioeconomic and healthcare variables The spread of COVID-19 was correlated negatively with aging index and positively with the other indicators showed.

Table 3 shows the bivariate and multivariate analyses, demonstrating that the spread of COVID-19 in the regional setting considered was negatively associated (p-value=0.003) with the population aging index, i.e. the ratio of the number of people aged 65 or more to the number of people aged 0-14 years multiplied by 100. It was positively associated with: public transportation per capita, i.e. the ratio of the yearly number of public transport passengers to the total population (p-value=0.012). It was also positively associated, albeit to a lesser extent (p-value=0.070), with the % of private long-term care hospital beds, and more strongly with the % of private acute care hospital beds (p-value=0.006).

**Table 3:**
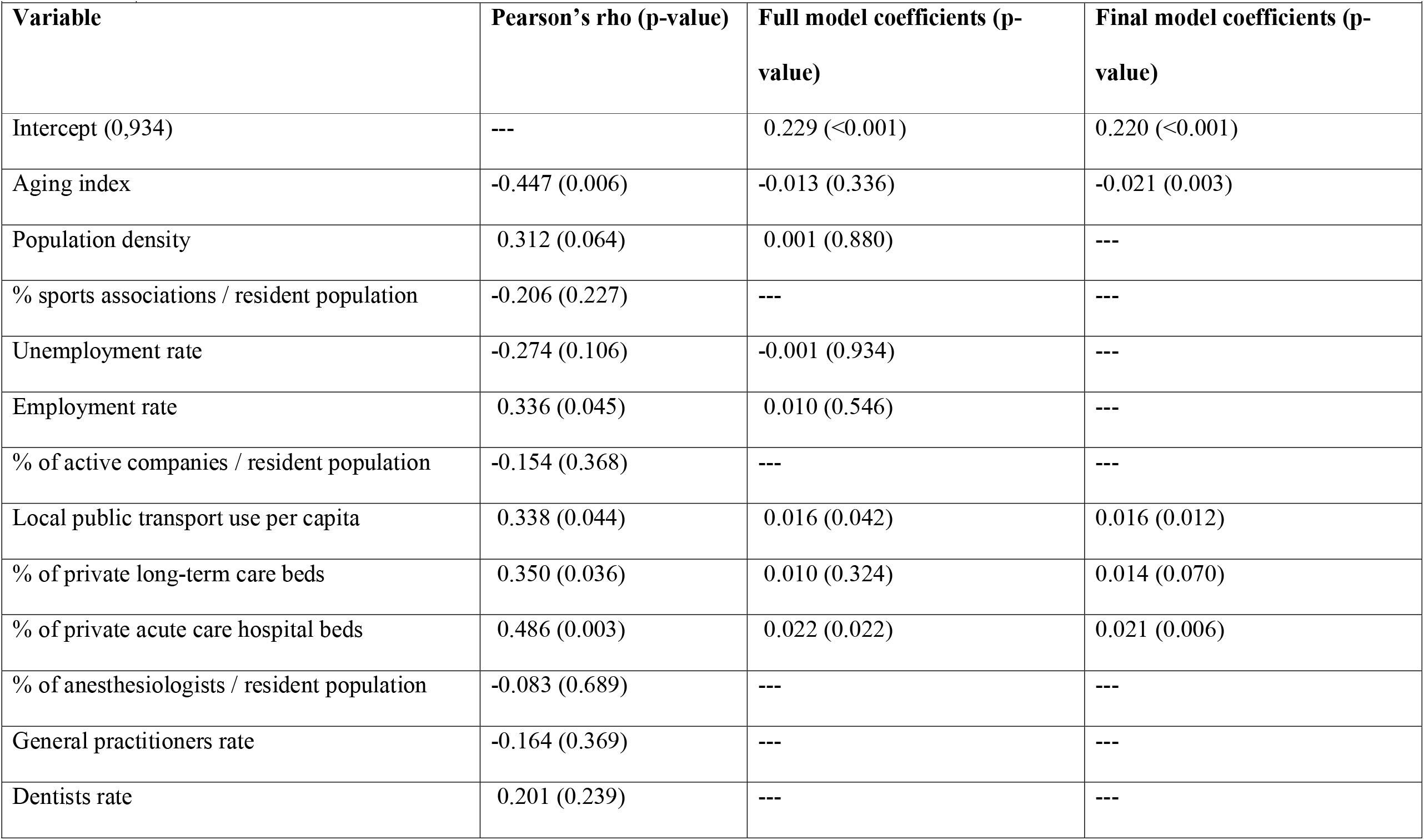
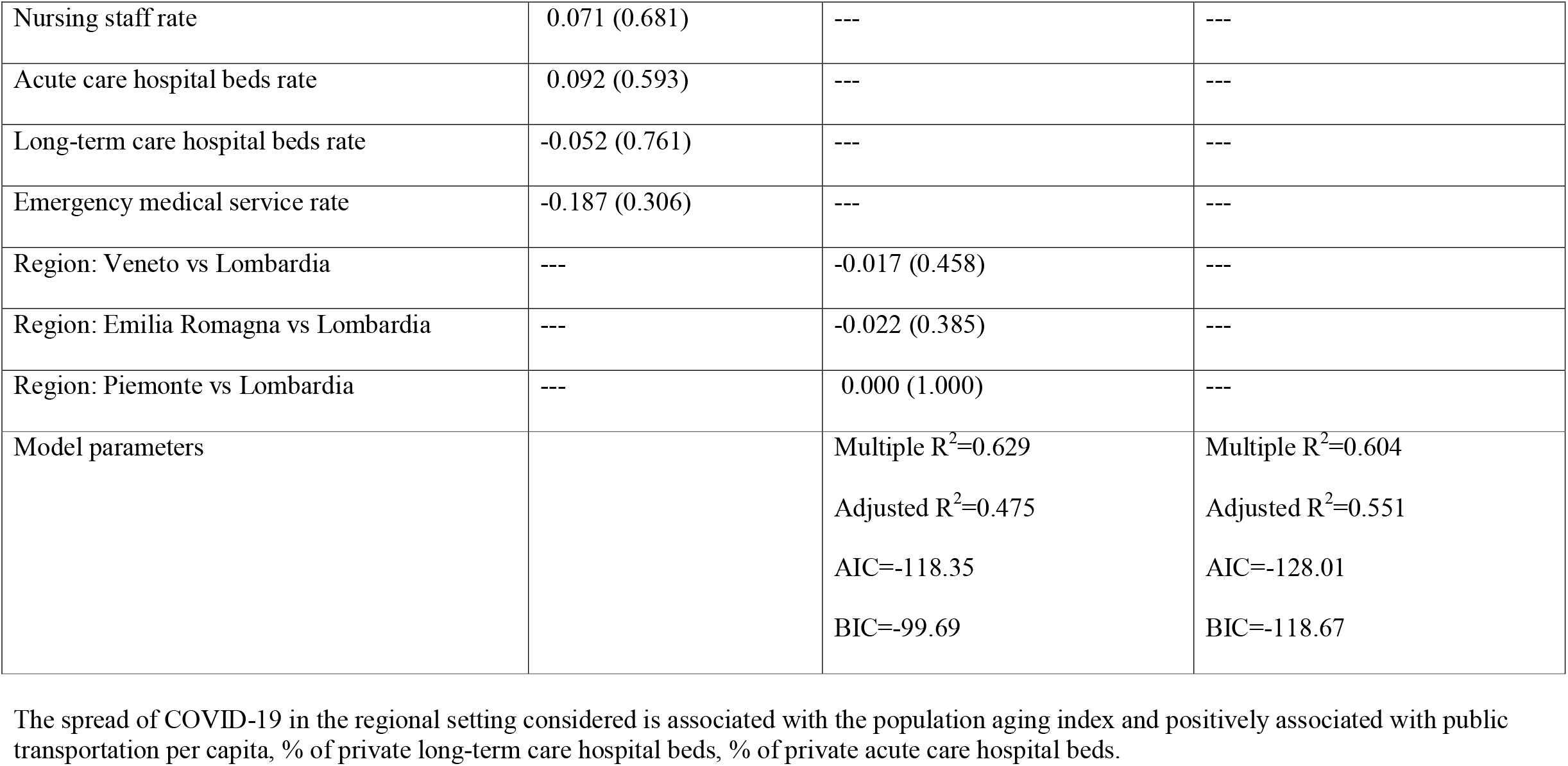
Association of covariates with COVID-19 transmission factor.

The Shapiro-Wilk test showed that the residuals had a normal distribution (p-value=0.870) and the Rubin-Watson test showed that the residuals were independent (p-value=0.48). The high value of the adjusted R squared indicates that the model had a good fit.

## DISCUSSION

With this ecological study, we found correlations between higher SARS-CoV-2 contagion rates and certain demographic, socioeconomic, and healthcare factors in the Italian provinces considered.

Unlike the first cases imported from outbreak regions, the local spread of communicable respiratory diseases seems to favor certain patterns and conditions, especially relating to social contact patterns.^13^

We found a negative association between COVID-19 contagion rates and aging. This seems somewhat at odds with the currently-known epidemiology of the virus, that in Italy is more likely to infect and kill older people. The median age of people infected with SARS-CoV-2 nationwide is 62 years old, and the median age of the related deaths is 80 years old.^14^ On the other hand, provinces with a higher aging index had a more gentle contagion curve. A possible explanation is that older people tend to move less outside their home and travel less far from where they live.^15^ They are also less likely to take part in mass gatherings or social events, whereas younger people tend to spend more time with others outside their family.^16^ Taken together, these behavioral factors could predispose the elderly to social distancing and self-isolation, naturally containing the propagation of communicable diseases in the provinces with the oldest populations.

In our analysis, higher levels of employment, public transportation usage, in-house density, and population density correlated positively with the spread of infection. What these socioeconomic factors have in common is the mobility of individuals and their exposure to close social contacts, both of which facilitate the propagation of SARS-CoV-2. This is consistent with recent analyses that associated a higher risk of COVID-19 transmission in mainland China with the use of trains,^17^ buses, and flights from Wuhan, and the risk was higher the longer a journey lasted.^18^ In-house density has also already been identified as a factor predisposing to higher transmission rates of pandemic influenza in Great Britain.^19^

A greater spread of the virus was seen in provinces with larger proportions of private acute care beds in clinics, and long-term care beds in nursing homes. These data should not be interpreted as a lack of involvement of the private sector in providing hospital care for COVID-19 patients. These facilities are an important part of the services provided by the Italian health system and they responded promptly to the emergency situation. They have ordinary and intensive care beds for COVID-19 patients just like public hospitals. The explanation should be sought by looking at the issue more in depth and speculating on the outcome variable analyzed in the present study, i.e. the rate of spread of the infection. Hospitals, clinics, and care homes are only responsible for containing it within their walls, while the preventive medicine departments of local health units are in charge of wider containment efforts, based on epidemiological intelligence.^20^ It may be that provinces with higher proportions of private healthcare facilities experienced more communication difficulties with the reporting of cases to preventive medicine departments to gather epidemic intelligence because private facilities are usually less integrated with preventive medicine and community healthcare services than public hospitals. This could hamper the early identification of patients through the sort of epidemic intelligence activities strongly recommended by the WHO in the case of COVID-19.^21^ Another explanation could relate to how private clinics are funded, i.e. based on DRG-based tariffs for each patient admitted. Owing to incentives linked to this particular funding mechanism, provinces with a relatively high concentration of private hospital beds might have paid less attention to avoiding the hospitalization of patients with COVID-19 symptoms who could have been successfully treated at home with the involvement of community and primary care services. In fact, primary healthcare services have a crucial role in ensuring that people caring for a family member suffering from COVID-19 manage their contact with the patient appropriately, and follow national or local policies regarding home quarantine. Caregivers should wear medical masks or the best available protection against respiratory droplets when in close contact with the patient, and observe hand hygiene recommendations. The WHO emphasized the importance of notifying healthcare providers of the diagnosis in order to receive instructions on where to seek care, when and where to enter healthcare facilities, and what precautions to take to prevent and control the infection.^21^ Our findings would suggest the importance of improving communications between private clinics and the Italian NHS responsible for their accreditation (i.e. the issue of the public license to provide hospital care on the NHS’s behalf) to ensure a real consistency of their activities with regional health plans (including their integration with the preventive medicine and primary care services of local health units), and an effective assessment of their activities and results.

This study has at least two important limitations. First, we collected data from available health indicators, which do not measure all the phenomena relevant to understanding and explaining the findings, even though our models performed well in explaining their variability (MR^2^ = ~60%). Second, the real number of SARS-CoV-2 contagions is known to be underestimated because not all individuals in the population considered were screened, and swabs were handled differently by the various regional healthcare systems. Even with such well-known limitations,^22^ ecological studies can still help healthcare workers and stakeholders to contain infections and fight pandemics, especially in the early stages of emerging diseases when clinical data are still limited.^23^

## Conclusions

Several demographic, socioeconomic, and healthcare factors were found associated with significant differences in the rate of COVID-19 spread in 36 provinces of Northern Italy. An older population seemed to naturally slow the contagion due to fewer social contacts. Socioeconomic factors (especially the rates of employment and public transportation usage) and organizational features of local healthcare systems (particularly the proportion of private healthcare facilities) also seemed to play an important part in the infection’s spread. These correlations could be helpful for the purpose of designing measures to reduce the propagation of COVID-19 in other countries now dealing with the first phase of this pandemic.

## Data Availability

Data are from publicly available repositories.

